# Sex differences in the coherence of PCL-5 and CAPS-5 assessments for Post-Traumatic Stress Disorder

**DOI:** 10.1101/2024.12.31.24319829

**Authors:** Bhavya Bakshi, Rebecca Nalloor, Vaughn McCall, Almira Vazdarjanova

## Abstract

The clinician-administered Post Traumatic Stress Disorder (PTSD) scale (CAPS) is a clinical tool for the diagnosis and assessment of the severity of PTSD symptoms. It is widely accepted as the gold standard against which self-assessment tools, such as PCL, are usually evaluated. However, the clinician administered interviews are time and resource intensive, thus utilizing self-report measures is adopted as quicker and more practical to assess symptom severity. Although there have been demonstrations of the validity of PCL for PTSD diagnosis and assessment of symptom severity, PCL (including PCL-5 which is based on DSM-5 criteria) scores tend to overreport symptom severity sometimes leading to higher prevalence of PTSD diagnosis. The contributing factors for such discrepancies in reporting are poorly understood. Here we examined whether there are sex differences in the alignment of PCL-5 with CAPS-5. Our findings indicate that males often report higher PTSD symptom severity on PCL-5 which significantly deviates from a predicted correlation line based on their CAPS-5 scores. Female participants had no deviation from the predicted correlation line. Accordingly, 26.8% of males and only 11.4% of females had inaccurate PTSD status if they were evaluated on PCL-5 alone. Our data also shows that participants with military trauma have poor correlation when compared to participants with civilian trauma. These findings have important implications for appropriate PTSD symptom severity assessment especially for males and military populations. This has significant downstream implications as treatment adjustments, therapy recommendations, and disease management, relying heavily on symptom severity assessed by self-reported tools.

## Background

Posttraumatic stress disorder (PTSD) symptoms are assessed using both structured interviews and self-reported measures (Weathers et al., 2013a, Weather el. 2013b). The clinician administered interviews are typically considered gold-standard (Weather et al., 2001) because interviewers can clarify responses, ask follow-up questions, and score severity of symptoms appropriately. However, the clinician administered interviews are time and resource intensive, thus utilizing self-report measures is quicker and more practical to assess symptom severity. Self-reported measures have great utility in practice; however, it is important to understand how self-reported measures and clinical interviews compare (Bovin & Weather, 2022).

The Clinician-Administered PTSD Scale (CAPS) is the gold standard clinician interview and the PTSD Checklist (PCL) is the most commonly used self-reported measure for PTSD symptom severity assessment (Weathers et al. 2013 a, b). Previous versions of CAPS and PCL had correlation ranges of 0.30 to 0.93 (Adkins et al., 2008; Blanchard et al., 1996; Bollinger et al., 2008; Forbes et al., 2001; Keen et al., 2008; Forbes et al, 2001). The latest versions of CAPS (CAPS-5; Weathers et al., 2018) and PCL (PCL-5; Belvins et al., 2015) which are based on DSM-5 (APA, 2013), aimed to increase alignment of scores. However, while both scales have been shown to be sound psychometric tools, higher PCL-5 symptom severity scores (31-33) compared to CAPS-5 (23, plus diagnostic criteria met) were reported to be optimally efficient for diagnosing PTSD (Bovin et al., 2016; Lee et al., 2022; Krüger-Gottschalk et al., 2022).

The difference in diagnostic scores between PCL-5 and CAPS-5 do not appear to result from differences in the range of the scales, study parameters, protocols, and goals. In military and veteran populations, not only did PCL-5 scores average 14 points higher, but PTSD symptoms were rated more severely and had a more evenly distributed score compared to CAPS-5’s bimodal distribution (Ressick et al, 2023). The study suggested that the self-reported nature of PCL-5 might lead to freer reporting or misinterpretation, resulting in higher scores, though it did not address the correlation between PCL-5 and CAPS-5. Furthermore, when PCL-5 and CAPS-5 scores from 378 veterans were standardized using Graphical Gaussian Models, PCL-5 reported higher symptom severity, although both tests shared similar symptom networks, indicating consistent symptom reporting (Moshier et al, 2018). It was suggested that PCL-5 and CAPS-5 predicted PTSD diagnoses with similar sensitivity and specificity. Similarly, Kruger-Gottschalk et al. (2017) validated the German version of PCL-5, demonstrating its high correlations of severity scores with CAPS-5, though their study did not focus on score differences. However, some research highlights the discrepancies across individual symptom scores measured by both tests (Kramer et al, 2023) and notes that PCL-5 yields higher prevalence estimation compared to a clinician administered test such as CAPS-5 (Marmar et al, 2015). Such evidence highlights the need for further investigation of the factors that drive test discrepancies to increase assessment reliability.

A possible contributor to the observed differences in PCL-5 and CAPS-5 could arise from differences in the participant populations such as differences in reporting between males and females. The purpose of this paper was to determine if sex differences affect the coherence between PCL-5 and CAPS-5 scores when reporting PTSD symptom severity. Our analyses also examined the predictive potential of PCL-5 for CAPS-5, deviations from the ideal correlation, and the effect of trauma type (military vs civilian).

## Methods

### Participants

The study included 100 adult (age ≥21 yrs old) male and female participants. Veteran status (Veteran vs Non-Veteran) and type of trauma (military vs civilian) were recorded because PTSD symptomatology differences have been reported between civilian and military populations. Each participant went through a CAPS-5 assessment with a trained interviewer and a PCL-5 self-assessment. Nineteen participants were from a previously published study which did not report the correlation between their PCL-5 and CAPS-5 scores (McCall et al 2018).

All participants in the study provided written, informed consent. The study was approved by the Research and Development Committee at VA Augusta Healthcare system and the IRB of record at Augusta University.

### Procedures and Measures

PCL-5 and CAPS-5 assessments were administered to each participant, in this order, to avoid possible interview influence on the self-reported measure.

CAPS-5 is a structured interview used to assess the presence and severity of Post-Traumatic Stress Disorder (PTSD) symptoms as defined by the Diagnostic and Statistical Manual of Mental Disorders, Fifth Edition (DSM-5). It is considered the gold standard for PTSD diagnosis and assessment. CAPS-5 assessed PTSD symptoms over the past month (Weathers et al., 2018). It was administered by a clinically trained interviewer and typically took 45-60 minutes to complete. The interview was administered only to those participants who met Criterion A (Index trauma) and after the participants had completed the PCL-5 assessment. CAPS-5 covered 30 items including:

1. Twenty DSM-5 PTSD symptoms that are grouped into four clusters (Cluster B: Intrusion, Cluster C: Avoidance, Cluster D: Negative alterations in cognitions and mood, and Cluster E: Arousal and reactivity).
2. Seven symptoms associated with PTSD (e.g., distress, functional impairment).
3. Three additional items assessing the onset and duration of symptoms, subjective distress, and functional impairment.

Each symptom was rated based on frequency and intensity on a 5-point scale from 0 to 4. The symptom severity scores were calculated by summing the frequency and intensity scores (0-8 for each symptom). Participants who met the DSM-5 PTSD criteria and had scores of greater than or equal to 23 were deemed to be PTSD (+), those with scores of less than 10 were PTSD (-), and the values in between were classified as subthreshold PTSD.

PCL-5 is a self-report questionnaire designed to assess the presence and severity of PTSD symptoms as defined by DSM-5. The PCL-5 questionnaire consists of 20 items that correspond to the 20 DSM-5 symptoms of PTSD. The items are grouped into the four DSM-5 symptom clusters, Cluster B-E. Participants individually completed the PCL-5 questionnaire which assessed symptoms over the past month. Each item on the PCL-5 was rated on a 5-point scale going from 0 to 4 and corresponding to degree of distress the respondent had experienced due to a particular symptom. The total score was calculated by summing the scores of all 20 items, resulting in a range from 0 to 80. Higher scores indicate more severe PTSD symptoms. For this study we used a total score of 34 or greater to classify a participant as PTSD (+). This score is commonly used a cut-off point for probable PTSD diagnosis (Bovin et al., 2016). Participants with values between 31 and 33 were deemed to be subthreshold PTSD and those with values of less than 31 were considered to be PTSD (-).

## Data Analysis

To examine if differences in CAPS-5 score existed between participant groups, we performed ANOVA tests with sex (males vs females) and trauma type (military vs civilian) as the independent variables.

The relationship between CAPS-5 (independent variable) and PCL-5 (dependent variable) was evaluated with a Spearman correlation analysis. To observe the correlation by sex, correlations were assessed for the entire data set (male and females combined), as well as males and females separately. Additionally, an odds ratio was calculated to assess the predictive value of PCL-5 for CAPS-5 within these groups.

To identify participants who may have overreported or underreported on the PCL-5 relative to their CAPS-5 score, we generated an ideal correlation line based on two intercepts: (0,0) for both PCL-5 and CAPS-5 and their respective PTSD threshold cutoffs (34 for PCL-5, 23 for CAPS-5). We analyzed potential sex differences in the deviation from the ideal correlation. First, we assessed the degree of deviation from the ideal line for males and females separately. This involved comparing the slopes and intercepts of the ideal correlation with the observed correlation using a t-test. Then, we compared the regression lines between males and females: regression coefficients (R^2^) for both groups were converted into a Z-scores with Fisher z transformation and we calculated a combined Z-score: z(obs)=(z(M)-z(F))/SqRt((1/(N(M)-3)+(1/(N(F)-3))) which determined whether the two regression lines were significantly different from each other. P-values of <0.05 were considered statistically significant.

Lastly, we analyzed the potential effect of trauma type—military versus civilian—on the correlation between CAPS-5 and PCL-5 across all participants, as well as males and females separately, using Spearman correlation.

## Results

A total of 100 participants, consisting of 56 males and 44 females, completed both the PCL-5 and CAPS-5 assessments. The participants had an average age of 39.8 years (males: 44.8 years, females: 33.4 years), with an average PCL-5 score of 42 (males: 45.7, females: 36.5) and an average CAPS-5 score of 24 (males: 23.1, females: 24.7).

### CAPS-5 scores were similar between male and female participants, and between military and civilian trauma participants

The CAPS-5 score was not statistically different between males and females (F(1,98)= 0.237, p = 0.6277) indicating that both groups had similar overall symptom severity. Additionally, no differences were observed in CAPS-5 score for military and civilian trauma (F(1,98)= 0.744, p = 0.3904).

### Female participants had a stronger correlation between self-reported and clinically assessed PTSD symptom scores

The correlation between PCL-5 and CAPS-5 scores was statistically significant for all participants (Rho = 0.815, p < 0.0001). However, when analyzed by sex, female participants exhibited a stronger correlation (Rho = 0.907, p < 0.0001) (Fig 1, magenta line) compared to male participants (Rho = 0.655, p < 0.0001) (Fig 2, blue line). The odds ratio for PCL-5 predicting CAPS-5 was markedly higher for females (144.6, p = 0.0010) than for males (11.8, p = 0.0031).

**Figure 1:**
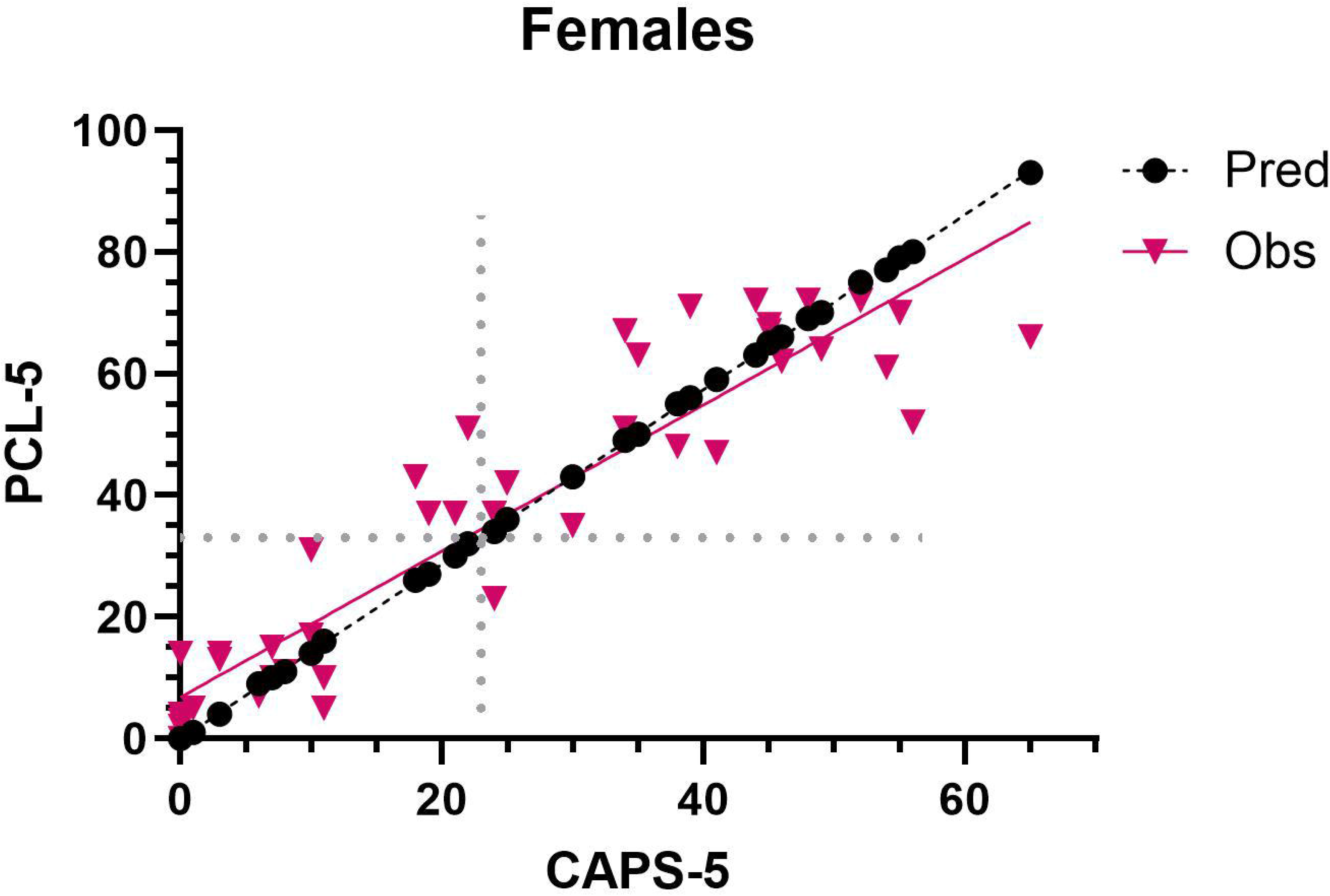
Correlation between CAPS-5 and PCL-5 symptom severity scores in **female participants**. Each point represents a participant. Magenta points are the reported/observed scores, while black points are those predicted based on the observed CAPS-5 scores. The grey lines are the CAPS-5 and PCL-5 cut-off values for PTSD diagnosis, such that top right and bottom left are those with coherent diagnoses. Participants with scores in the top left would be classified as PTSD by PCL-5 but not by CAPS-5. Those in the bottom right would be classified as noPTSD by PCL-5 but PTSD by CAPS-5.

**Figure 2:**
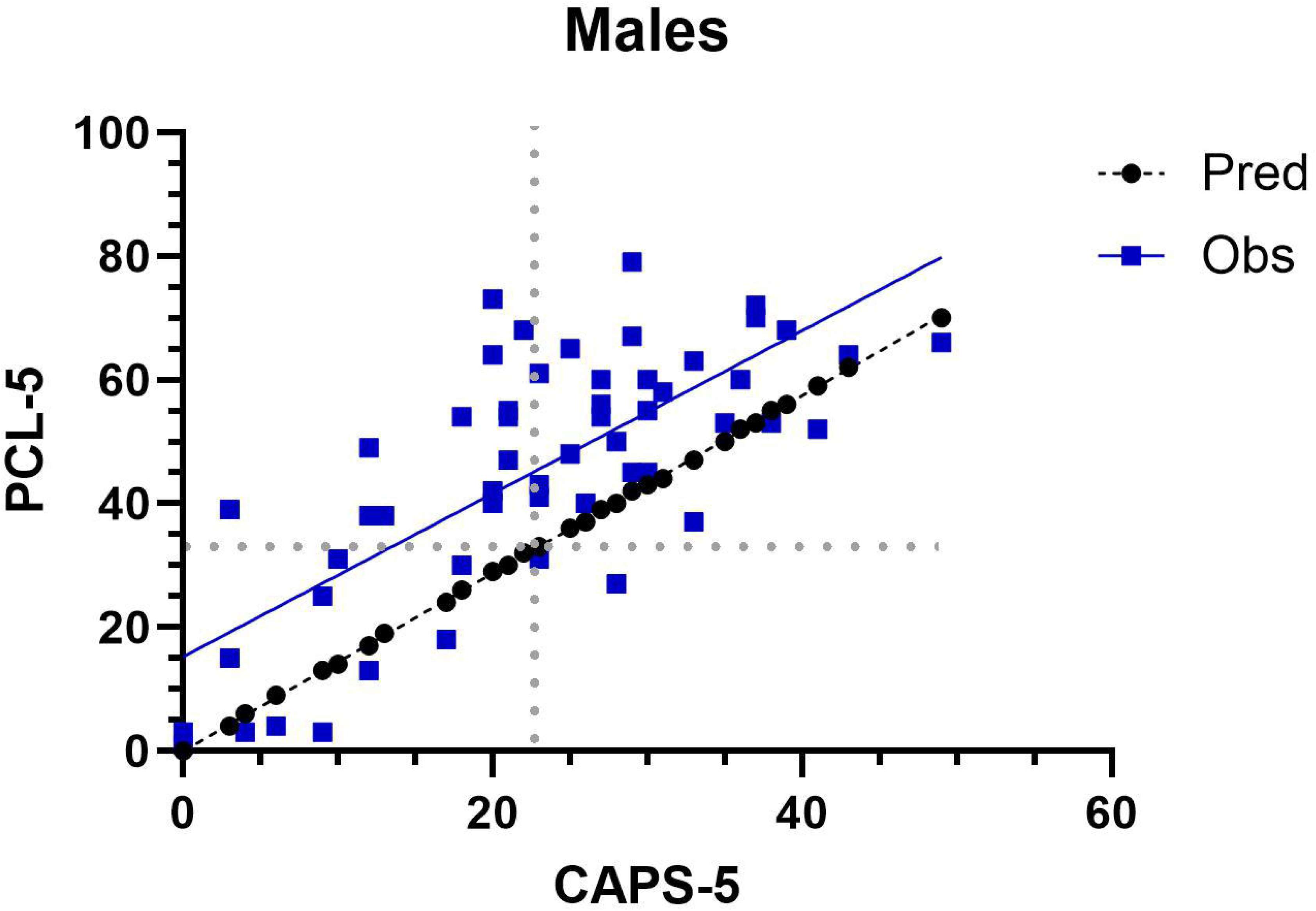
Correlation between CAPS-5 and PCL-5 symptom severity scores in **male participants**. Each point represents a participant. Blue points are the reported/observed scores, while black points are those predicted based on the observed CAPS-5 scores. The grey lines are the CAPS-5 and PCL-5 cut-off values for PTSD diagnosis, such that top right and bottom left are those with coherent diagnoses. Participants with scores in the top left would be classified as PTSD by PCL-5 but not by CAPS-5. Those in the bottom right would be classified as noPTSD by PCL-5 but PTSD by CAPS-5.

### Only male participants had significant deviation from the ideal/predicted regression line

For male participants, the observed values significantly deviated from the ideal regression line (Fig 2, black line), with a mean difference of 12.5 (p = 0.001) as determined by a paired t-test (Fig 4). This was not the case for female participants (Fig 1, black line), where the mean difference was minimal (1.068, p = 0.5077, Fig 3), indicating no significant deviation from the ideal regression line. The ideal regression was based on a PCL-5 score of 33 correlating with a CAPS-5 score of 23, representing thresholds for PTSD diagnosis. Additionally, a significant sex difference was observed in the deviation from the ideal correlation line between PCL-5 and CAPS-5 scores (Z(Observed) = -3.5, p= 0.0005). Combined, these findings suggest that females have a stronger correlation between self-reported and clinically assessed PTSD symptoms.

**Figure 3:**
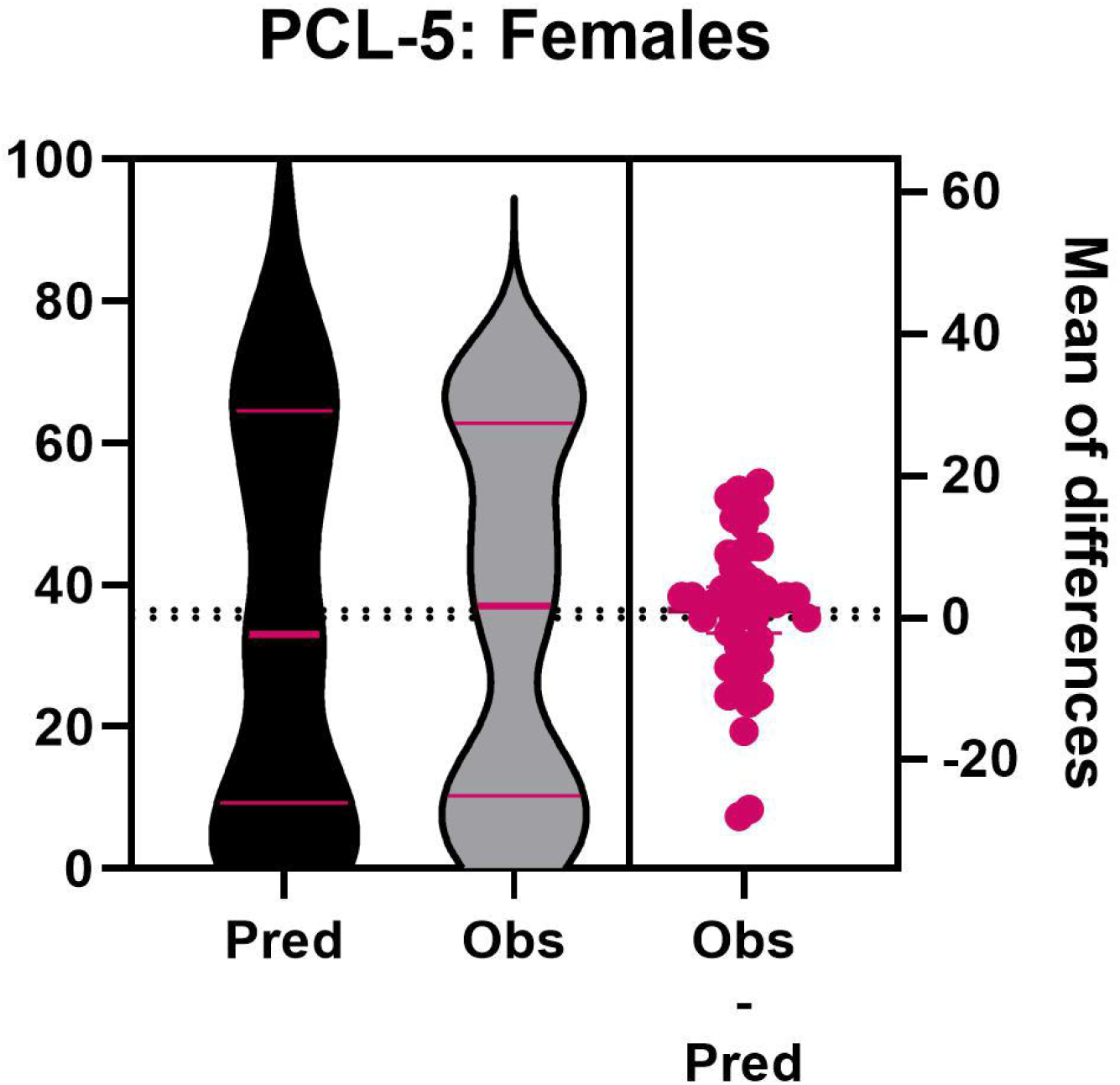
Distribution of predicted (based on CAPS-5 scores) and observed PCL-5 symptom severity scores in **female participants**.

**Figure 4:**
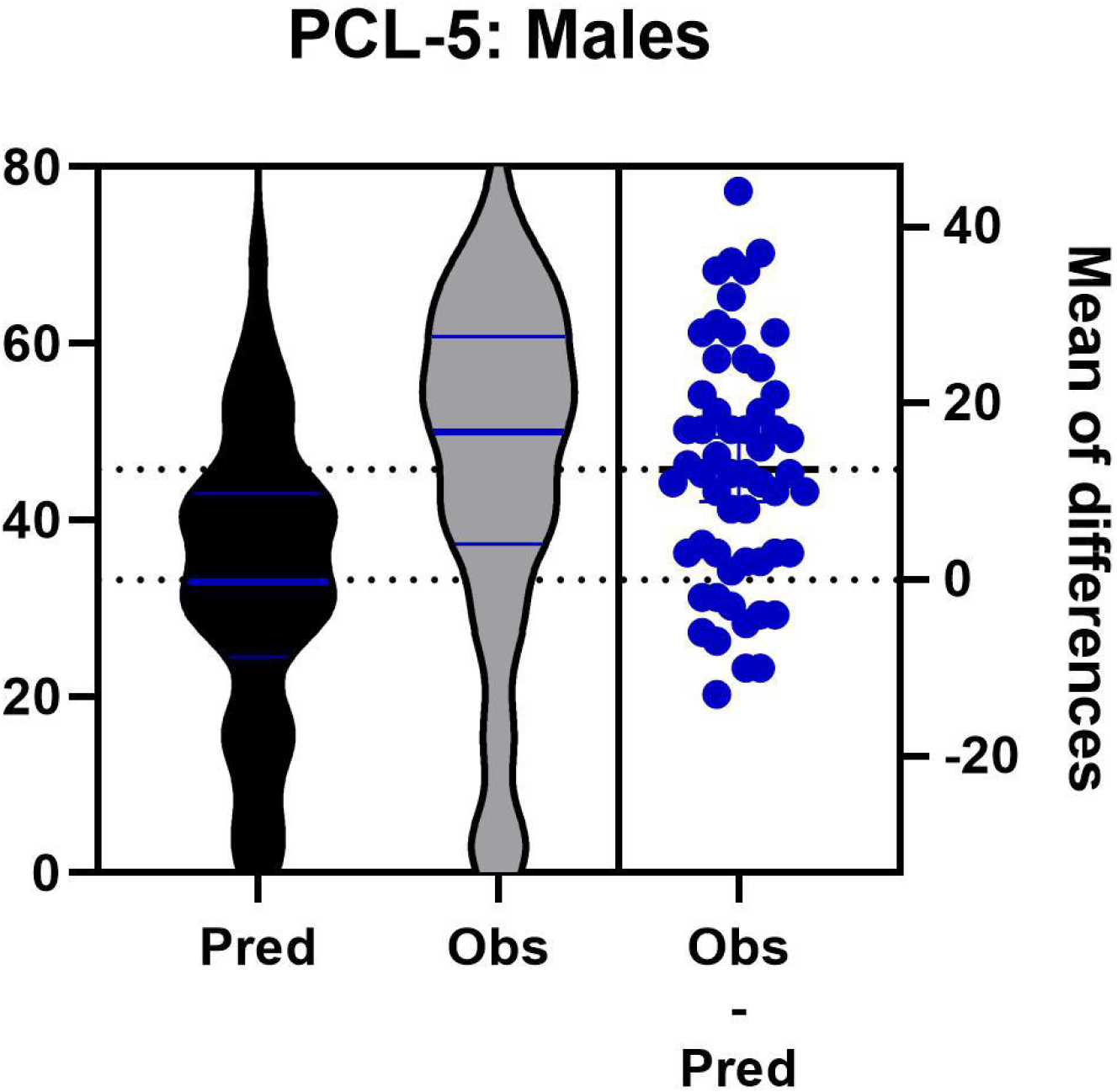
Distribution of predicted (based on CAPS-5 scores) and observed PCL-5 symptom severity scores in **male participants**.

### Males had a higher proportion of misclassified PTSD cases compared to females based on PCL-5 alone

Among males, a total of 15 (26.8%) were misclassified if PCL-5 scores were used compared to CAPS-5 scores: 13 (23.2%) participants had PCL-5 scores that met the PTSD+ criteria but failed to meet the criteria on CAPS-5 (Fig 2, top left quadrant).

Conversely, 2 (3.6%) males were PTSD-per PCL-5 scores, yet their CAPS-5 scores and assessment met the criteria for PTSD+ (Fig 2, bottom right quadrant).

In contrast, only 5 (11.4%) of the female participants were misclassified: 4 (9.1%) participants were PTSD+ on PCL-5 while not meeting the criteria on CAPS-5 (PTSD-) (Fig 1, top left quadrant). Only 1 (2.8%) female participant was found to be PTSD-on PCL-5 but PTSD+ on CAPS-5 (Fig 1, bottom right quadrant). These findings are consistent with the strengths of the odds ratios reported above.

Combined, these observations suggest that the weaker correlation between PCL-5 and CAPS-5 scores in males resulted in a higher proportion of misclassified PTSD cases compared to females. These findings are consistent with the observed left shift of the PCL-5/CAPS-5 regression line compared to the ideal regression line in males but not females (Figs 1&2).

### Civilian trauma had stronger correlation than military trauma

When considering trauma type, the correlation between PCL-5 and CAPS-5 was stronger among participants with civilian trauma (Rho = 0.885, p < 0.0001, n = 62) than those with military trauma (Rho = 0.523, p = 0.0015, n = 38). This pattern held true within each sex. Additionally, a significant sex difference was observed in the correlations between civilian trauma and military trauma for PCL-5 and CAPS-5 (Z(Observed) = 7.26, p= 0.0005).

Males with civilian trauma had a stronger correlation (Rho = 0.801, p = 0.0002, n = 22), while those with military trauma had a weaker correlation (Rho = 0.470, p = 0.0070, n = 34), despite a higher sample in the latter. This appeared to be the case for females as well: females with civilian trauma showed a stronger correlation (Rho = 0.903, p < 0.0001, n = 39) compared to military trauma (Rho = 0.600, p = 0.2301, n = 4), although the sample size in the military group was too small to warrant a strong inference.

## Discussion

This study examined the relationship between self-reported PCL-5 and clinician administered CAPS-5 assessments for PTSD symptom severity across male and female participants. Firstly, CAPS-5 scores had no difference between male and female participants indicating equivalent PTSD symptom severity status for the participants. The study revealed significant sex differences in the correlation between PCL-5 and CAPS-5. Notably, female participants exhibited a stronger correlation between PCL-5 and CAPS-5 compared to their male counterparts. This finding suggests that females may report their symptoms more consistently across self-reported and clinician administered measures.

One possible explanation for this discrepancy could be that males may inaccurately self-report their PTSD symptoms due to external factors such as differences in symptom presentation. For instance, males may experience more irritability and reckless behavior while females may experience more avoidance and hyperarousal (Green 2003, Fullerton et al., 2001). Additional external factors could include secondary gain associated with PTSD diagnosis, overidentification with trauma symptoms, and societal expectations that could discourage men from expressing emotional distress. Conversely, females are probably more likely to accurately self-report their PTSD symptoms due more willingness to seek healthcare in general (Bertakis et al., 2000) and lower stigma around obtaining care for mental health.

We also tested a practical implication of the discrepancy in PCL-5/CAPS-5 reporting and assessment, respectively: The higher discrepancy in males, compared to females, will result in a higher proportion of male participants with misclassified PTSD status according to PCL-5 vs CAPS-5 assessment. The results supported this hypothesis. It should be stressed that the severity of symptoms was similar between males and females. This misclassification trend highlights the potential limitations of relying solely on self-reported measures for diagnosing PTSD, particularly in male populations. The discrepancy between self-reported and clinically assessed PTSD symptoms could result in over- or under-diagnosis, leading to inappropriate or inadequate treatment. These considerations are significant considering that more than a quarter of our male participants were misclassified. Our findings are consistent with reported findings of higher PCL-5 scores than CAPS-5 scores for participants (Bovin et al., 2016); Lee et al., 2022); Krüger-Gottschalk et al., 2022). However, no other studies isolated for sex differences, and noted changes in the magnitude of discrepancy between PCL-5 and CAPS-5 based on sex. The existing studies reported populations with a high severity of symptoms have a greater chance at being assessed appropriately by PCL-5, and populations with borderline PTSD presentation, PCL-5 requires cut-off manipulation for appropriate categorization (Bovin and Marx, 2023). It is possible that the utility of PCL-5 for clinical PTSD diagnosis decreases as the intensity of the symptoms decreases. A larger study capturing the continuity of symptoms with sufficient sample size along the continuum is needed to test this hypothesis.

Examining the differences between the ideal and observed regressions further highlights the differences between male and female participants. Males had a high mean difference and a significant deviation of the ideal vs observed regression lines. In contrast, females had minimal deviation and a non-significant low mean difference. This finding further suggests that PCL-5 may be less reliable at predicting PTSD symptom severity for males as their self-reported scores were significantly different from expected values. Additionally, the mean difference of 12.5 is high in magnitude and has implications of misclassifying participants, as already discussed above.

The study also examined the effect of trauma type (civilian vs. military) on the correlation between PCL-5 and CAPS-5 scores and whether the effect is similar for both sexes. The results indicated that civilian trauma was associated with a stronger correlation compared to military trauma across all participants. We further showed that this pattern was true for both sexes. This is consistent with previously reported differences in trauma type affecting PTSD symptomology, treatment outcomes, and clinical presentation (Guina et al., 2018; Jakob et al., 2017; Kessler et al., 2017). However, no specific study has looked at PCL-5 and CAPS-5 correlation based on trauma type.

These findings suggest that the nature of trauma may influence how individuals perceive and report PTSD symptoms and how accurately these reports align with clinical assessments. The weaker correlation in military trauma cases, particularly among males, could reflect the unique psychological and social factors associated with military service, such as stigma surrounding mental health issues or the impact of combat experiences on symptom expression (Guina et al., 2018; Jakob et al., 2017; Kessler et al., 2017).

The results of this study have important implications that can impact clinical practice in the future, thus require further examination. First, it is important to consider sex and trauma type as factors when assessing PTSD symptoms with a self-reported measure like PCL-5. For males, additional caution may be needed when interpreting the PCL-5 self-reported measure as it may not fully capture PTSD symptom severity and lead to misclassification. Incorporating additional assessment tools could help mitigate the risk and improve appropriate symptom severity monitoring.

## Limitations

This study provides valuable insights into the relationship between PCL-5 and CAPS-5 across sexes and trauma types. However, several limitations should be acknowledged. Firstly, the sample size of 100 participants is not very large, affecting generalizability of the findings. For instance, female participants with military trauma were few. Secondly, participants were only from the southeast region and therefore could contribute geographical effects that we were unable to account for. By considering these limitations, future research can be designed to address these gaps, leading to more comprehensive and generalizable findings on the relationship between PCL-5 and CAPS-5 across different populations and contexts.

## Conclusion

This study highlighted sex differences in the correlation of self-reported PCL-5 and clinician-administered CAPS-5 for PTSD assessment. Although these measures were correlated in both sexes, females showed stronger alignment between them. This discrepancy in alignment was associated with a higher number of male participants with misclassified PTSD status. The findings also feature the importance of considering trauma type (civilian vs. military) when assessing PTSD, particularly in male and military populations. The results indicate further work is needed to establish the causation of these discrepancies and caution clinicians to be mindful of these differences.

## Data Availability

All data produced in the present study are available upon reasonable request to the authors

## Author contributions

BB wrote the manuscript with input from AV, VM and RN. AV and RN participated in the designing of the experiments. RN, VM and AV collected the data. AV and BB independently conducted data analyses.

## Acknowledgements

This work was supported by the Department of Veterans Affairs, Veterans Health Administration, Office of Research and Development, Biomedical Laboratory Research and Development, I01BX001978 and I01BX003890.

The contents of this manuscript do not represent the views of the VA or the United States Government.

## References

1. Adkins, J. W., Weathers, F. W., McDevitt-Murphy, M., & Daniels, J. B. (2008). Psychometric properties of seven self-report measures of posttraumatic stress disorder in college students with mixed civilian trauma exposure. Journal of Anxiety Disorders, 22(8), 1393–1402. 10.1016/j.janxdis.2008.02.002 [PubMed] [CrossRef] [Google Scholar] [Ref list]

2. American Psychiatric Association. (2013). Diagnostic and Statistical Manual of Mental Disorders (5th ed.). 10.1176/appi.books.9780890425596 [CrossRef] [Google Scholar] [Ref list]

3. Bertakis, K. D., Azari, R., Helms, L. J., Callahan, E. J., & Robbins, J. A. (2000). Gender differences in the utilization of health care services. The Journal of family practice, 49(2), 147–152.

4. Blanchard, E. B., Jones Alexander, J., Buckley, T. C., & Forneris, C. A. (1996). Psychometric properties of the PTSD Checklist (PCL). Behaviour Research and Therapy, 34(8), 669–673. 10.1016/0005-7967(96)00033-2 [PubMed] [CrossRef] [Google Scholar] [Ref list]

5. Blevins, C. A., Weathers, F. W., Davis, M. T., Witte, T. K., & Domino, J. L. (2015). The Posttraumatic Stress Disorder Checklist for DSM-5 (PCL-5): development and initial psychometric evaluation. Journal of Traumatic Stress, 28(6), 489–498. 10.1002/jts.22059 [PubMed] [CrossRef] [Google Scholar] [Ref list]

6. Bollinger, A. R., Cuevas, C. A., Vielhauer, M. J., Morgan, E. E., & Keane, T. M. (2008). The operating characteristics of the PTSD Checklist in detecting PTSD in HIV+ substance abusers. Journal of Psychological Trauma, 7(4), 213–234. 10.1080/19322880802384251 [CrossRef] [Google Scholar] [Ref list]

7. Bovin, M. J., & Weathers, F. W. (2022). Assessing PTSD symptoms. In Beck G. & Sloan D. (Eds.), Oxford Handbook of Traumatic Stress Disorders (2nd ed, pp. 235–249). Oxford University Press. [Google Scholar] [Ref list]

8. Bovin, M. J., Marx, B. P., Weathers, F. W., Gallagher, M. W., Rodriguez, P., Schnurr, P. P., & Keane, T. M. (2016). Psychometric properties of the PTSD Checklist for Diagnostic and Statistical Manual of Mental Disorders Fifth Edition (PCL-5) in veterans. Psychological Assessment, 28(11), 1379–1391. 10.1037/pas0000254 [PubMed] [CrossRef] [Google Scholar] [Ref list]

9. Bovin, M. J., Marx, B. P., Weathers, F. W., Gallagher, M. W., Rodriguez, P., Schnurr, P. P., & Keane, T. M. (2016b). Psychometric properties of the PTSD Checklist for Diagnostic and Statistical Manual of Mental Disorders–Fifth Edition (PCL-5) in veterans. Psychological Assessment, 28(11), 1379–1391. 10.1037/pas0000254

10. Bovin, Michelle J, and Brian P Marx. “The Problem with Overreliance on the PCL–5 as a Measure of PTSD Diagnostic Status.” Clinical Psychology-Science and Practice, vol. 30, no. 1, 1 Mar. 2023, pp. 122–125, 10.1037/cps0000119.

11. Forbes, D., Creamer, M., & Biddle, D. (2001). The validity of the PTSD Checklist as a measure of symptomatic change in combat-related PTSD. Behaviour Research and Therapy, 39(8), 977–986. 10.1016/S0005-7967(00)00084-X [PubMed] [CrossRef] [Google Scholar] [Ref list]

12. Forbes, D., Creamer, M., & Biddle, D. (2001). The validity of the PTSD Checklist as a measure of symptomatic change in combat-related PTSD. Behaviour Research and Therapy, 39(8), 977–986. 10.1016/S0005-7967(00)00084-X [PubMed] [CrossRef] [Google Scholar] [Ref list]

13. Fullerton, C. S., Ursano, R. J., Epstein, R. S., Crowley, B., Vance, K., Kao, T. C., Dougall, C., & Baum, A. (2001). Gender differences in posttraumatic stress disorder after motor vehicle accidents. American Journal of Psychiatry, 158(9), 1486–1491.

14. Green, B. (2003). Post-traumatic stress disorder: Symptom profiles in men and women. Current Medical Research and Opinion, 19(3), 200–204. 10.1185/030079903125001604

15. Guina, J., Nahhas, R. W., Sutton, P., & Farnsworth, S. (2018). The Influence of Trauma Type and Timing on PTSD Symptoms. The Journal of nervous and mental disease, 206(1), 72–76. 10.1097/NMD.0000000000000730

16. Jakob, J. M., Lamp, K., Rauch, S. A., Smith, E. R., & Buchholz, K. R. (2017). The Impact of Trauma Type or Number of Traumatic Events on PTSD Diagnosis and Symptom Severity in Treatment Seeking Veterans. The Journal of nervous and mental disease, 205(2), 83–86. 10.1097/NMD.0000000000000581

17. Keen, S. M., Kutter, C. J., Niles, B. L., & Krinsley, K. E. (2008). Psychometric properties of PTSD checklist in sample of male veterans. Journal of Rehabilitation Research & Development, 45(3), 465–474. 10.1682/JRRD.2007.09.0138 [PubMed] [CrossRef] [Google Scholar] [Ref list]

18. Kessler, R. C., Aguilar-Gaxiola, S., Alonso, J., Benjet, C., Bromet, E. J., Cardoso, G., Degenhardt, L., de Girolamo, G., Dinolova, R. V., Ferry, F., Florescu, S., Gureje, O., Haro, J. M., Huang, Y., Karam, E. G., Kawakami, N., Lee, S., Lepine, J. P., Levinson, D., Navarro-Mateu, F., … Koenen, K. C. (2017). Trauma and PTSD in the WHO World Mental Health Surveys. European journal of psychotraumatology, 8(Sup5), 1353383. 10.1080/20008198.2017.1353383

19. Kramer, L. B., Whiteman, S. E., Petri, J. M., Spitzer, E. G., & Weathers, F. W. (2023). Self-Rated Versus Clinician-Rated Assessment of Posttraumatic Stress Disorder: An Evaluation of Discrepancies Between the PTSD Checklist for DSM-5 and the Clinician-Administered PTSD Scale for DSM-5. Assessment, 30(5), 1590–1605. 10.1177/10731911221113571

20. Krüger-Gottschalk, A., Ehring, T., Knaevelsrud, C., Dyer, A., Schäfer, I., Schellong, J., Rau, H., & Köhler, K. (2022). Confirmatory factor analysis of the clinician-administered PTSD scale (CAPS-5) based on DSM-5 vs. ICD-11 criteria. European Journal of Psychotraumatology, 13(1), 2010995. 10.1080/20008198.2021.2010995 [PMC free article] [PubMed] [CrossRef] [Google Scholar] [Ref list]

21. Lee, D. J., Weathers, F. W., Thompson-Hollands, J., Sloan, D. M., & Marx, B. P. (2022). Concordance in PTSD symptom change between DSM-5 versions of the Clinician-Administered PTSD Scale and the PTSD Checklist (PCL-5). Psychological Assessment, 34(6), 604–609. 10.1037/pas0001130 [PMC free article] [PubMed] [CrossRef] [Google Scholar] [Ref list]

22. Livingston, N. A., Brief, D. J., Miller, M. W., & Keane, T. M. (2021). Assessment of PTSD and its comorbidities in adults chapter 16. In Friedman M.J., Schnurr P.P., & Keane T.M. (Eds.), Handbook of PTSD: Science and practice (3rd ed., pp. 283–298). The Guilford Press. [Google Scholar] [Ref list]

23. Marmar, C. R., Schlenger, W., Henn-Haase, C., Qian, M., Purchia, E., Li, M., Corry, N., Williams, C. S., Ho, C.-L., Horesh, D., Karstoft, K. I., Shalev, A., & Kulka, R. A. (2015). Course of posttraumatic stress disorder 40 years after the Vietnam War: Findings from the National Vietnam Veterans Longitudinal Study. Journal of the American Medical Association Psychiatry, 72, 875–881. 10.1001/jamapsy-chiatry.2015.0803

24. McCall, W. V., Pillai, A., Case, D., McCloud, L., Nolla, T., Branch, F., Youssef, N. A., Moraczewski, J., Tauhidul, L., Pandya, C. D., & Rosenquist, P. B. (2018). A Pilot, Randomized Clinical Trial of Bedtime Doses of Prazosin Versus Placebo in Suicidal Posttraumatic Stress Disorder Patients With Nightmares. Journal of clinical psychopharmacology, 38(6), 618–621. 10.1097/JCP.0000000000000968

25. Moshier, S. J., Bovin, M. J., Gay, N. G., Wisco, B. E., Mitchell, K. S., Lee, D. J., Sloan, D. M., Weathers, F. W., Schnurr, P. P., Keane, T. M., & Marx, B. P. (2018). Examination of posttraumatic stress disorder symptom networks using clinician-rated and patient-rated data. J Abnorm Psychol, 127(6), 541–547. 10.1037/abn0000368

26. Resick, P. A., Straud, C. L., Wachen, J. S., LoSavio, S. T., Peterson, A. L., McGeary, D. D., Young-McCaughan, S., Taylor, D. J., Mintz, J., Consortium, S. S., & the Consortium to Alleviate, P. (2023). A comparison of the CAPS-5 and PCL-5 to assess PTSD in military and veteran treatment-seeking samples. Eur J Psychotraumatol, 14(2), 2222608. 10.1080/20008066.2023.2222608

27. Weathers, F. W., Blake, D. D., Schnurr, P. P., Kaloupek, D. G., Marx, B. P., & Keane, T. M. (2013a). The clinician-administered PTSD scale for DSM-5 (CAPS-5) [Assessment]. U.S. Department of Veterans Affairs, National Center for PTSD. https://www.ptsd.va.gov/professional/assessment/adult-int/caps.asp. [Ref list]

28. Weathers, F. W., Bovin, M. J., Lee, D. J., Sloan, D. M., Schnurr, P. P., Kaloupek, D. G., Keane, T. M., & Marx, B. P. (2018). The clinician-administered PTSD scale for DSM–5 (CAPS-5): development and initial psychometric evaluation in military veterans. Psychological Assessment, 30(3), 383–395. 10.1037/pas0000486 [PMC free article] [PubMed] [CrossRef] [Google Scholar] [Ref list]

29. Weathers, F. W., Litz, B. T., Keane, T. M., Palmieri, P. A., Marx, B. P., & Schnurr, P. P. (2013b). The PTSD checklist for DSM-5 (PCL-5) [Assessment]. U.S. Department of Veterans Affairs, National Center for PTSD. https://www.ptsd.va.gov/professional/assessment/adult-sr/ptsd-checklist.asp. [Ref list]

30. Weathers, F.W., Blake, D.D., Schnurr, P.P., Kaloupek, D.G., Marx, B.P., & Keane, T.M. (2013). The Clinician-Administered PTSD Scale for DSM-5 (CAPS-5).

